# Revisiting the Area Deprivation Index

**DOI:** 10.64898/2026.02.26.26346490

**Authors:** Keying Chen, Bradley G. Hammill

## Abstract

**Objective:** To re-estimate and re-validate the Area Deprivation Index to address recent criticism of the existing index, which is calculated and distributed by Neighborhood Atlas.

**Data Sources:** To calculate the updated Area Deprivation Index (ADI), we obtained 17 census measures from the 2018–2022 American Community Survey (ACS) 5-year data that reflected poverty, housing, employment, and education within census block groups, census tracts, and counties. To validate the association of the updated index with health outcomes, we obtained the most recent census tract-level life table data from the U.S. Small-area Life Expectancy Estimates Project (USALEEP).

**Study Design:** We used principal components analysis to estimate new factor weights associated with the primary component summarizing the 17 selected census measures. The resulting scores were ranked into percentiles to arrive at the updated ADI. We compared this updated ADI to the existing ADI distributed by the Neighborhood Atlas. We calculated average life expectancy and age-specific mortality rates by groups defined by the updated ADI.

**Principal Findings:** The principal components analysis identified a single factor that explained 46% of variance at the census tract level. There were noticeable differences in factor loadings for the unemployment rate and the proportion of households without a motor vehicle compared to the original loadings. Results were similar at the county and block group levels. Compared to existing ADI values, there were substantive changes in the updated ADI values for many geographic ares with high home values, but low employment and educational attainment. The updated ADI demonstrated robust associations with age-specific mortality and life expectancy.

**Conclusions:** The updated ADI better summarized the 17 underlying census variables than the current ADI. The updated index was strongly correlated with life expectancy and mortality.

**Callout Box:** What is known on this topic

- ADI is a measure of area-level deprivation that summarizes 17 Census measures regarding poverty, housing, employment, and education. The original ADI demonstrated associations with mortality and clinical outcomes.
- The current version of the ADI made available by the Neighborhood Atlas relies on old factor weights and appears to have flaws in its construction that lead to overreliance on home value and income variables.

What this study adds

- To address these concerns, this study re-estimates the ADI properly from source data to ensure that the final index reflects a combination of all 17 census measures, and not just home value and income.
- The updated ADI more accurately reflects the distribution of deprivation in neighborhoods across the country and is highly correlated with life expectancy and age-specific mortality rates.
- The updated ADI is available publicly and should be used by researchers who would like to utilize a broad measure of neighborhood deprivation.

## Introduction

The Area Deprivation Index (ADI) was introduced in 2003 (Singh) as a summary measure of area-level deprivation, based on 17 variables from 1990 Census data that reflected different aspects of poverty, housing, employment, and education.^1^ At the county level, this original index demonstrated robust associations with all-cause mortality rates at all ages in the United States.^1^ Based on this work, the Neighborhood Atlas was introduced in 2018 to make the ADI more widely available for research.^2^ The Neighborhood Atlas uses data from the American Community Survey (ACS) to calculate the index and keep it up to date.

In recent years, however, multiple studies have reported that the Neighborhood Atlas ADI is overwhelmingly associated with median home value and median income, resulting in the situation where cities with high home values had no neighborhoods that appeared to be deprived, contradicting the accepted understanding of the distribution of deprivation within these cities.^3,4^ These studies strongly suggested that the calculation of the ADI by the Neighborhood Atlas was performed improperly, leading variables with naturally larger scales to dominate the index, undermining the ADI’s ability to accurately reflect broad-based area-level deprivation. Because programming code and raw measure scores are not made available by the Neighborhood Atlas, it is not possible to definitively confirm this suspicion.

In response, recommendations have been put forth to improve the accuracy and transparency of any social deprivation measure.^5^ These include calls to provide clear details about index construction methods, to make measure code publicly available, and to regularly update and revalidate the measure. In this study, we address these suggestions by calculating the ADI from source ACS data (2018-2022), using principal components analysis to estimate updated factor loadings; by validating the updated ADI by estimating associations with mortality and clinical outcomes; and by making our code and results publicly available.

## Methods

### Estimating the ADI

The American Community Survey (ACS) is an ongoing survey conducted annually by the U.S. Census Bureau, collecting data from over 3.5 million households across the country.^6^ The ACS provides information regarding social, economic, housing, and demographic characteristics, and can be used to allocate resources, assess community needs, and guide policy decisions at local, state, and national level. We used the ACS 5-year data from 2018 to 2022 at the census block group, census tract, and county level.^7^

With two minor changes, we used the same 17 ACS-based measures regarding poverty, housing, employment, and education used in the original ADI.^1,8^ These measures are listed in **Table 1**, along with the ACS source table on which they were based and the calculation used, based on the variables in those tables. The first change we made was to replace the discontinued “Households without a telephone” measure with the “Households without an internet connection” measure. The second change we made was to update the household income thresholds used in the calculation of the income disparity variable to reflect the general increase in incomes over time. The original index defined income disparity based on the ratio of the number of households with <$10,000 income to number of households with ≥$50,000 income.^1^ In 1990, there were 9.0% of households with <$10,000 income and 31.0% of households with ≥$50,000 income, while in the 2018–2022 ACS data the percentages based on these thresholds were 4.9% and 66.0%, respectively.^9,10^ We updated these thresholds to $20,000 and $125,000, which yielded similar proportions in each tail to the 1990 data. We retained a variable based on the original thresholds to understand the effect that not updating them has on the resulting index.

**Table 1.** List and source of variables used for calculation of the Area Deprivation Index.

| Measure | ACS/Census Table | Variable Definition |
| --- | --- | --- |
| **Poverty** |  |  |
| Median family income, \$ | B19113 | B19113_001 |
| Income disparity | B19001 | Using original income cut-offs: $B19001\_E002 / \text{sum}(B19001\_E011, B19001\_E012, B19001\_E013, B19001\_E014, B19001\_E015, B19001\_E016, B19001\_E017)$<br>Using updated income cut-offs: $\text{sum}(B19001\_E002, B19001\_E003, B19001\_E004) / \text{sum}(B19001\_E015, B19001\_E016, B19001\_E017)$ |
| Families below poverty level, % | B17010 | $B17010\_E002 / B17010\_E001$ |
| Population below 150% poverty threshold, % | C17002 | $\text{sum}(C17002\_E002, C17002\_E003, C17002\_E004, C17002\_E005) / C17002\_E001$ |
| Single parent households with dependents <18, % | B11005 | $B11005\_E005 / B11005\_E001$ |
| Households without a motor vehicle, % | B25044 | $\text{sum}(B25044\_E003, B25044\_E010) / B25044\_E001$ |
| Households without an internet connection, % | B28002 | $B28002\_E013 / B28002\_E001$ |
| Occupied housing units without complete plumbing, % | B25049 | $(B25049\_E004 + B25049\_E007) / B25049\_E001$ |
| **Housing** |  |  |
| Owner occupied housing units, % | B25003 | $B25003\_E002 / B25003\_E001$ |
| Households with >1 person per room, % | B25014 | $\text{sum}(B25014\_E005, B25014\_E006, B25014\_E007, B25014\_E011, B25014\_E012, B25014\_E013) / B25014\_E001$ |
| Median monthly mortgage, \$ | B25088 | B25088_E001 |
| Median gross rent, \$ | B25064 | B25064_E001 |
| Median home value, \$ | B25077 | B25077_E001 |
| **Employment** |  |  |
| Employed person 16+ in white collar occupation, % | C24010 | $\text{sum}(C24010\_E003, C24010\_E027, C24010\_E039, C24010\_E063) / C24010\_E001$ |
| % |  |  |
| Civilian labor force unemployed (aged 16+), % | B23025 | B23025_E005 / B23025_E003 |
| **Education** |  |  |
| Population aged 25+ with <9yr education, % | B15003 | sum(B15003_E002, B15003_E003, B15003_E004, B15003_E005, B15003_E006, B15003_E007, B15003_E008, B15003_E009, B15003_E010, B15003_E011, B15003_E012) / B15003_E001 |
| Population aged 25+ with at least a high school education, % | B15003 | sum(B15003_E017, B15003_E018, B15003_E019, B15003_E020, B15003_E021, B15003_E022, B15003_E023, B15003_E024, B15003_E025) / B15003_E001 |
| **Exclusion criteria** |  |  |
| Number of persons | B99011 | B99011_E001 |
| Number of households | B19001 | B19001_E001 |
| Population living in group quarters, % | P17 | P17_029N / P17_001N |

All tables listed above, except P17, are 2022 American Community Survey (ACS) 5-year summary files. Data and documentation for the ACS files can be found at https://www.census.gov/programs-surveys/acs/data/summary-file.html. The U.S. Census P17 table, which can be found at https://data.census.gov/table/DECENNIALDHC2020.P17, was used for information about group quarters population. All variable names reflect the field names assigned in the downloadable files.

Finally, since most of the 17 measures included in the ADI have skewed distributions, we created a parallel set of variables that were transformed for normality using log, square root, or square functions, as appropriate. Even though having normally-distributed inputs are not required for principal components analysis (PCA),^11^ we were interested in exploring the effect that such transformations would have on the resulting index.

We used PCA to reduce the dimensionality of the 17 source measures, while retaining as much information as possible, at the county, census tract, and census block group levels. In SAS, the PCA procedure itself normalizes the input variables to have mean equal to 0 and standard deviation equal to 1 prior to analysis.^12^ In R, this standardization often needs to be specified manually prior to or during the PCA.^13^ We summarize the PCA results by describing the structure of the identified principal components, the explained variance, the factor weightings, and the scoring coefficients. We compare our results at the census tract level to the PCA results reported by Singh.

The updated ADI was calculated as the percentiles (1 to 100) of the PCA-based score distribution. We report the correlations that the source measures have with the resulting ADI; and we compare the updated ADI values to ADI values from the Neighborhood Atlas for the same census block group.

### Validating the ADI

To validate the updated ADI, we took two approaches. First, to see if the updated ADI reflected our expectation of the geography of deprivation, we mapped ADI values for census block groups within both the District of Columbia and Manhattan. For comparison, we did the same for the ADI values from the Neighborhood Atlas. Second, we used life table data from the United States Small-Area Life Expectancy Estimates Project (USALEEP) available from the Centers for Disease Control and Prevention (CDC) to explore the relationship, at the census tract level, between the updated ADI deciles and life expectancy at birth (*e*_0_), life expectancy at other ages (*e_x_*), and annualized age-specific mortality (_1_*m_x_*).^14^

All analyses were performed in parallel in both SAS (v9.4) and R (v4.4.0).^15,16^ The data used for these analyses are all publicly available. We made our programming code, analysis output, and updated ADI values available publicly at { https://gitlab.oit.duke.edu/duke-adi/2022}.

The Duke Health Institutional Review Board approved this study (Pro00106448).

## Results

Summary statistics for the 17 source measures at the census tract level are presented in **Table 2**. Most of the source measures exhibited considerable skewness in their untransformed distributions. Following application of appropriate transformations, the skewness coefficients improved substantially across nearly all measures. There were only modest amounts of missing data among census tracts (**Table 3**), primarily for financial measures—median family income (1.2%) and median gross rent (4.7%)—which were imputed using county-level data. At the census block group level, financial variables showed higher rates of missing data, with median gross rent missing most frequently (>27%). Most of this missing data was able to be imputed using census tract information.

**Table 2.** Summary statistics for source measures at the census tract level.

| Measure | Mean (SD) | Min–Max | Skewness Coefficient, (untransformed variable) | Skewness Coefficient (transformed variable) | Transformation applied |
| --- | --- | --- | --- | --- | --- |
| **Poverty** |  |  |  |  |  |
| Median family income, \$ | \$96,177 (\$45,079) | \$2,499–\$250,001 | 1.18 | –0.30 | Natural log |
| Income disparity<sup>a</sup> | 2.17 (6.63) | 0.02–50.00 | 5.90 | 0.90 | Natural log |
| Families below poverty level, % | 10.2 (10.5) | 0–100 | 2.02 | 0.34 | Square root |
| Population below 150% poverty threshold, % | 22.1 (15.1) | 0–100 | 1.12 | 0.48 | Square root |
| Single parent households with dependents <18, % | 10.6 (7.6) | 0–100 | 1.37 | 0.30 | Square root |
| Households without a motor vehicle, % | 8.6 (11.7) | 0–98.2 | 3.20 | 0.12 | Square root |
| Households without an internet connection, % | 10.0 (8.5) | 0–93.7 | 1.64 | 1.20 | Square root |
| Occupied housing units without complete plumbing, % | 0.4 (1.3) | 0–53.9 | 10.51 | 0.18 | Square root |
| **Housing** |  |  |  |  |  |
| Owner occupied housing units, % | 65.1 (23.4) | 0–100 | –0.78 | –0.10 | Square |
| Households with >1 person per room, % | 3.6 (5.3) | 0–70.0 | 3.14 | 0.89 | Square root |
| Median monthly mortgage, \$ | \$1422 (\$758) | \$99–\$4001 | 1.05 | –0.30 | Natural log |
| Median gross rent, \$ | \$1344 (\$594) | \$99–\$3501 | 1.22 | 0.08 | Natural log |
| Median home value, \$ | \$358,759 (\$293,654) | \$9999–\$2,000,001 | 2.44 | 0.09 | Natural log |
| **Employment** |  |  |  |  |  |
| Employed person 16+ in white collar occupation, % | 59.7 (15.7) | 0–100 | 0.03 | –0.37 | Square root |
| Civilian labor force unemployed (aged 16+), % | 5.7 (4.6) | 0–71.9 | 2.22 | 0.43 | Square root |
| **Education** |  |  |  |  |  |
| Population aged 25+ with <9yr education, % | 5.0 (6.3) | 0–78.2 | 2.69 | 0.94 | Square root |
| Population aged 25+ with at least a high school education, % | 88.6 (9.9) | 9.3–100 | –1.68 | –1.23 | Square |

<sup>a</sup> We constrained this ratio to be between 0.02 and 50

**Table 3.** Imputation information for source measure missing data.

| Measure | Census tract-level measures<br>(N = 83,005) | Census block group-level measures<br>(N = 236,303) |  |
| --- | --- | --- | --- |
| Measure | Imputed w/county data<br>N (%) | Imputed w/tract data<br>N (%) | Imputed w/county data<br>N (%) |
| **Poverty** |  |  |  |
| Median family income, \$ | 980 (1.2) | 21,841 (9.2) | 1279 (0.5) |
| Income disparity | 0 (0.0) | 0 (0.0) | 0 (0.0) |
| Families below poverty level, % | 6 (<0.1) | 165 (<0.1) | 9 (<0.1) |
| Population below 150% poverty threshold, % | 0 (0.0) | 0 (0.0) | 0 (0.0) |
| Single parent households with dependents <18, % | 0 (0.0) | 0 (0.0) | 0 (0.0) |
| Households without a motor vehicle, % | 0 (0.0) | 0 (0.0) | 0 (0.0) |
| Households without an internet connection, % | 0 (0.0) | 0 (0.0) | 0 (0.0) |
| Occupied housing units without complete plumbing, % | 0 (0.0) | 0 (0.0) | 0 (0.0) |
| **Housing** |  |  |  |
| Owner occupied housing units, % | 0 (0.0) | 0 (0.0) | 0 (0.0) |
| Households with >1 person per room, % | 0 (0.0) | 0 (0.0) | 0 (0.0) |
| Median monthly mortgage, \$ | 1995 (2.4) | 21,245 (9.0) | 3726 (1.6) |
| Median gross rent, \$ | 3879 (4.7) | 55,860 (23.6) | 8276 (3.5) |
| Median home value, \$ | 1660 (2.0) | 16,090 (6.8) | 3105 (1.3) |
| **Employment** |  |  |  |
| Employed person 16+ in white collar occupation, % | 1 (<0.1) | 43 (<0.1) | 1 (<0.1) |
| Civilian labor force unemployed (aged 16+), % | 1 (<0.1) | 35 (<0.1) | 1 (<0.1) |
| Measure | Imputed w/county data<br>N (%) | Imputed w/tract data<br>N (%) | Imputed w/county data<br>N (%) |
| **Education** |  |  |  |
| Population aged 25+ with <9yr education, % | 0 (0.0) | 8 (<0.1) | 0 (0.0) |
| Population aged 25+ with at least a high school education, % | 0 (0.0) | 8 (<0.1) | 0 (0.0) |

The principal components analysis identified a single dominant factor that adequately summarized the 17 component measures across all geographic levels. At the census tract level, the first component explained 40% and 46% of the variance for untransformed and transformed variables, respectively (**Table 4**). The original ADI explained 52% of the variance. The improved explanatory power when using transformed variables demonstrates the value of addressing skewness in the source measures. The percentages of variance explained were marginally higher at the county level (**Table 5**), and marginally lower at the block group level (**Table 6**).

**Table 4.** Factor loadings for the updated ADI compared to the original ADI at the census tract level.

| Measure | Updated ADI (2018–2022 data) |  | Original ADI<br>(1990 data) |
| --- | --- | --- | --- |
|  | Untransformed<br>variables | Transformed<br>variables |  |
| Poverty |  |  |  |
| Median family income, \$ | -0.85 | -0.91 | -0.86 |
| Income disparity | 0.54 | 0.89 | 0.83 |
| Families below poverty level, % | 0.80 | 0.82 | 0.86 |
| Population below 150% poverty threshold, % | 0.89 | 0.90 | 0.92 |
| Single parent households with dependents <18, % | 0.66 | 0.63 | 0.63 |
| Households without a motor vehicle, % | 0.40 | 0.49 | 0.61 |
| Households without an internet connection, % | 0.69 | 0.73 | 0.77 |
| Occupied housing units without complete plumbing, % | 0.19 | 0.21 | 0.45 |
| Housing |  |  |  |
| Owner occupied housing units, % | -0.47 | -0.49 | -0.54 |
| Households with >1 person per room, % | 0.41 | 0.39 | 0.49 |
| Median monthly mortgage, \$ | -0.63 | -0.70 | -0.68 |
| Median gross rent, \$ | -0.60 | -0.64 | -0.69 |
| Median home value, \$ | -0.55 | -0.69 | -0.61 |
| Employment |  |  |  |
| Employed person 16+ in white collar occupation, % | -0.79 | -0.79 | -0.77 |
| Civilian labor force unemployed (aged 16+), % | 0.49 | 0.45 | 0.71 |
| Education |  |  |  |
| Population aged 25+ with <9yr education, % | 0.61 | 0.63 | 0.75 |
| Population aged 25+ with at least a high school education, % | -0.77 | -0.76 | -0.86 |
| Proportion of total variance explained | 0.40 | 0.46 | 0.52 |

**Table 5.** Factor loadings for the updated ADI compared to the original ADI at the county level.

| Measure | Updated ADI (2018–2022 data) |  | Original ADI<br>(1990 data) |
| --- | --- | --- | --- |
|  | Untransformed<br>variables | Transformed<br>variables |  |
| Poverty |  |  |  |
| Median family income, \$ | -0.90 | -0.94 | -0.92 |
| Income disparity | 0.61 | 0.93 | 0.88 |
| Families below poverty level, % | 0.87 | 0.89 | 0.88 |
| Population below 150% poverty threshold, % | 0.91 | 0.93 | 0.93 |
| Single parent households with dependents <18, % | 0.59 | 0.56 | 0.33 |
| Households without a motor vehicle, % | 0.43 | 0.51 | 0.45 |
| Households without an internet connection, % | 0.77 | 0.80 | 0.78 |
| Occupied housing units without complete plumbing, % | 0.27 | 0.31 | 0.64 |
| Housing |  |  |  |
| Owner occupied housing units, % | -0.14 | -0.16 | -0.44 |
| Households with >1 person per room, % | 0.27 | 0.22 | 0.40 |
| Median monthly mortgage, \$ | -0.77 | -0.85 | -0.78 |
| Median gross rent, \$ | -0.66 | -0.70 | -0.79 |
| Median home value, \$ | -0.64 | -0.74 | -0.67 |
| Employment |  |  |  |
| Employed person 16+ in white collar occupation, % | -0.63 | -0.63 | -0.69 |
| Civilian labor force unemployed (aged 16+), % | 0.59 | 0.54 | 0.57 |
| Education |  |  |  |
| Population aged 25+ with <9yr education, % | 0.65 | 0.66 | 0.79 |
| Population aged 25+ with at least a high school education, % | -0.78 | -0.79 | -0.82 |
| Proportion of total variance explained | 0.43 | 0.49 | 0.51 |

**Table 6.** Factor loadings for the updated ADI at the census block group level.

| Measure | Updated ADI (2018–2022 data) |  |
| --- | --- | --- |
|  | Untransformed variables | Transformed variables |
| **Poverty** |  |  |
| Median family income, \$ | -0.82 | -0.86 |
| Income disparity | 0.55 | 0.82 |
| Families below poverty level, % | 0.70 | 0.72 |
| Population below 150% poverty threshold, % | 0.83 | 0.85 |
| Single parent households with dependents <18, % | 0.54 | 0.51 |
| Households without a motor vehicle, % | 0.37 | 0.45 |
| Households without an internet connection, % | 0.59 | 0.63 |
| Occupied housing units without complete plumbing, % | 0.14 | 0.15 |
| **Housing** |  |  |
| Owner occupied housing units, % | -0.47 | -0.49 |
| Households with >1 person per room, % | 0.34 | 0.32 |
| Median monthly mortgage, \$ | -0.63 | -0.68 |
| Median gross rent, \$ | -0.59 | -0.62 |
| Median home value, \$ | -0.57 | -0.68 |
| **Employment** |  |  |
| Employed person 16+ in white collar occupation, % | -0.72 | -0.70 |
| Civilian labor force unemployed (aged 16+), % | 0.37 | 0.31 |
| **Education** |  |  |
| Population aged 25+ with <9yr education, % | 0.55 | 0.55 |
| Population aged 25+ with at least a high school education, % | -0.70 | -0.69 |
| **Proportion of total variance explained** | 0.34 | 0.39 |

Table 1 presents the factor loadings for the updated ADI, based on both transformed and untransformed source variables, at the census tract level. The factor loadings were largely similar between these different versions, with the largest differences for some of the financial variables. For income disparity, the transformed variable showed a substantially stronger loading (0.89) compared to the untransformed variable (0.54). These loadings are both much stronger than for the version of income disparity based on the original income thresholds (0.29, untransformed; 0.75, transformed). The transformed version of the updated ADI also showed stronger loadings for median home value (−0.69, transformed vs. −0.55, untransformed) and median monthly mortgage (–0.70 vs. –0.63).

There were several notable differences between our updated ADI version (transformed variables) and Singh’s original ADI (Table 1) at the census tract level. The civilian unemployment rate had a loading of 0.71 in the original ADI, but only a loading of 0.45 in the updated ADI. The percentage of housing without complete plumbing was less important in the updated ADI (loading, 0.21), compared to the original ADI (0.45). Other smaller differences in loadings were seen for motor vehicle ownership rates, household crowding, and the education variables.

Factor loadings for the two updated ADI versions and for the original ADI at the county level are shown in Table 5; and loadings for the two updated ADI versions at the block group level are shown in Supplemental Table 6. Singh did not report results at the block group level in the original ADI development. Except for slightly higher loading of unemployment rates and lower loadings of owner-occupied rates and household crowding and at the county level, compared to the census tract levels the results were largely similar at these geographies to the results from the census tract level.

The scoring coefficients for all geographic levels, for both transformed and untransformed source variables, are reported in **Table 7**, **Table 8**, and **Table 9**.

**Table 7.** Scoring coefficients for the updated ADI at the census tract level.

| Measure | Updated ADI (2018–2022 data) |  |
| --- | --- | --- |
|  | Untransformed variables | Transformed variables |
| **Poverty** |  |  |
| Median family income, \$ | -0.1243 | -0.1154 |
| Income disparity | 0.0793 | 0.1128 |
| Families below poverty level, % | 0.1178 | 0.1042 |
| Population below 150% poverty threshold, % | 0.1296 | 0.1148 |
| Single parent households with dependents <18, % | 0.0960 | 0.0798 |
| Households without a motor vehicle, % | 0.0590 | 0.0623 |
| Households without an internet connection, % | 0.1008 | 0.0929 |
| Occupied housing units without complete plumbing, % | 0.0285 | 0.0264 |
| **Housing** |  |  |
| Owner occupied housing units, % | -0.0684 | -0.0618 |
| Households with >1 person per room, % | 0.0594 | 0.0492 |
| Median monthly mortgage, \$ | -0.0928 | -0.0893 |
| Median gross rent, \$ | -0.0885 | -0.0811 |
| Median home value, \$ | -0.0810 | -0.0869 |
| **Employment** |  |  |
| Employed person 16+ in white collar occupation, % | -0.1157 | -0.1001 |
| Civilian labor force unemployed (aged 16+), % | 0.0724 | 0.0572 |
| **Education** |  |  |
| Population aged 25+ with <9yr education, % | 0.0891 | 0.0793 |
| Population aged 25+ with at least a high school education, % | -0.1122 | -0.0963 |

**Table 8.** Scoring coefficients for the updated ADI at the county level.

| Measure | Updated ADI (2018–2022 data) |  |
| --- | --- | --- |
|  | Untransformed variables | Transformed variables |
| **Poverty** |  |  |
| Median family income, \$ | -0.1237 | -0.1130 |
| Income disparity | 0.0835 | 0.1118 |
| Families below poverty level, % | 0.1195 | 0.1077 |
| Population below 150% poverty threshold, % | 0.1250 | 0.1122 |
| Single parent households with dependents <18, % | 0.0813 | 0.0675 |
| Households without a motor vehicle, % | 0.0593 | 0.0616 |
| Households without an internet connection, % | 0.1061 | 0.0971 |
| Occupied housing units without complete plumbing, % | 0.0365 | 0.0373 |
| **Housing** |  |  |
| Owner occupied housing units, % | -0.0186 | -0.0195 |
| Households with >1 person per room, % | 0.0365 | 0.0266 |
| Median monthly mortgage, \$ | -0.1048 | -0.1027 |
| Median gross rent, \$ | -0.0907 | -0.0843 |
| Median home value, \$ | -0.0883 | -0.0891 |
| **Employment** |  |  |
| Employed person 16+ in white collar occupation, % | -0.0859 | -0.0761 |
| Civilian labor force unemployed (aged 16+), % | 0.0814 | 0.0656 |
| **Education** |  |  |
| Population aged 25+ with <9yr education, % | 0.0895 | 0.0796 |
| Population aged 25+ with at least a high school education, % | -0.1068 | -0.0948 |

**Table 9.**
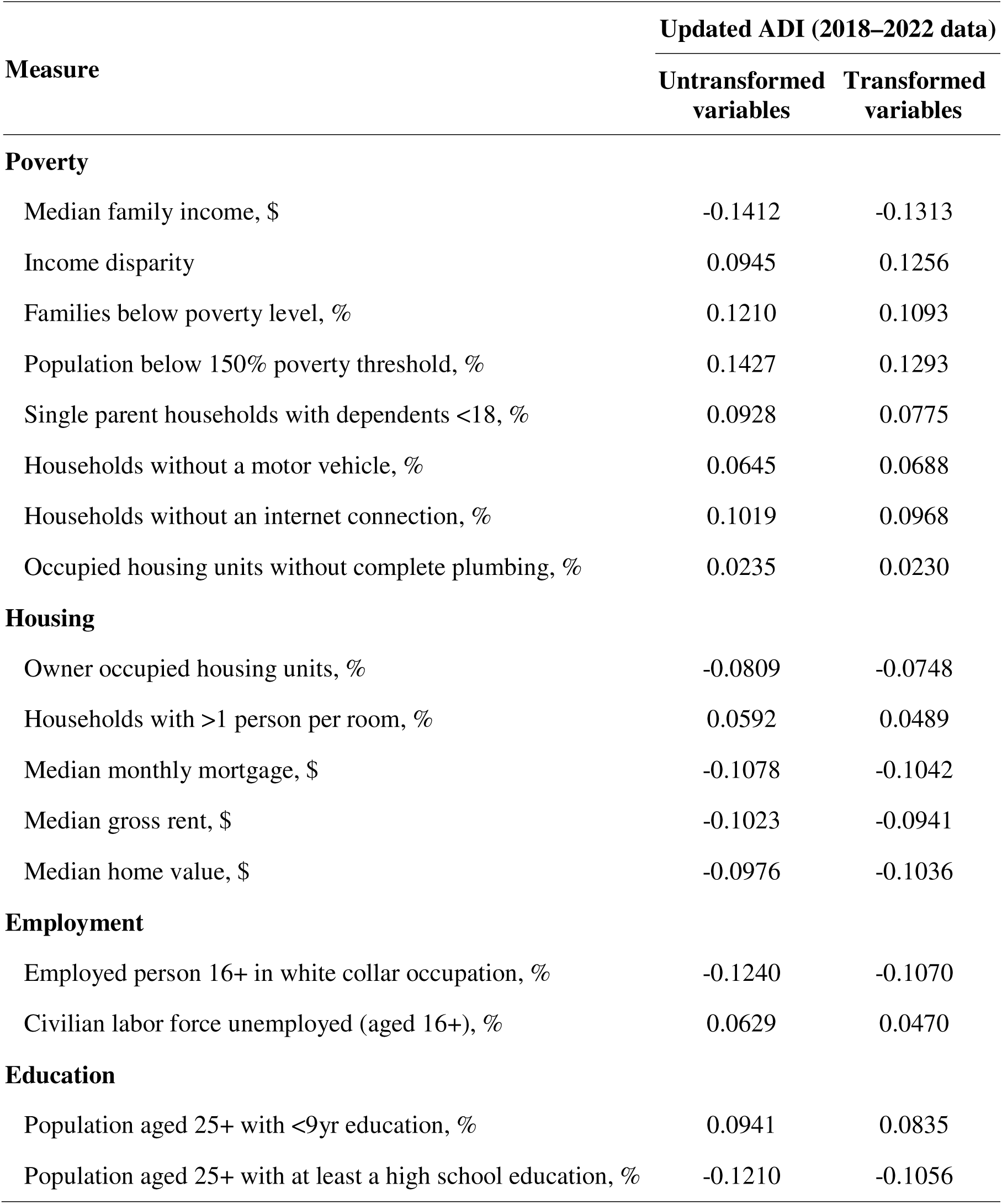
Scoring coefficients for the updated ADI at the census block group level.

The updated ADI (transformed variables) showed strong associations with life expectancy across the lifespan (**Figure 1**, **Table 10**). At birth, life expectancy differed by 7.6 years between census tracts in the lowest deprivation quintile (81.9 years) and those in the highest deprivation quintile (74.3 years). Life expectancy differences between these two groups only dropped below 5 years at age 55.

**Figure 1.**
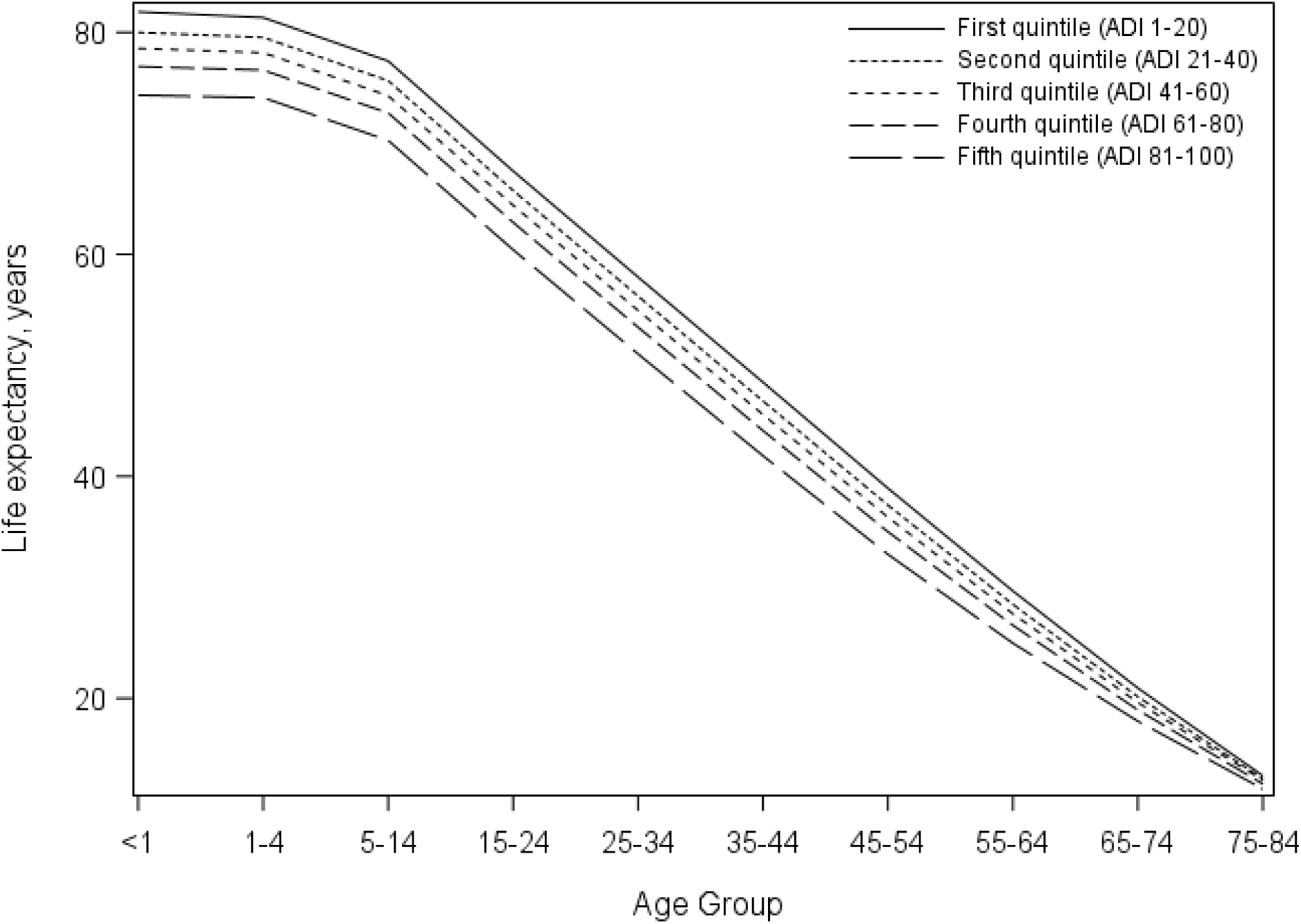
Life expectancy at different ages by quintile of the updated Area Deprivation Index at the census tract level. The first quintile of ADI (1-20) reflects the census tracts with the least deprivation while the fifth quintile of ADI (81-100) reflects the census tracts with the highest deprivation. The life expectancy associated with the first age category shown (<1 year) reflects the life expectancy at birth. *To change: Add “years” to the x-axis labels*

**Table 10.** Mean (SD) tract-level life expectancy, in years, by life table age group and quintiles of ADI.

| Life Table Age Group<sup>a</sup> | Quintile of Duke ADI |  |  |  |  | Difference in LE<br>Quintile 5 – Quintile 1 |
| --- | --- | --- | --- | --- | --- | --- |
|  | 1 | 2 | 3 | 4 | 5 |  |
| 0-1 years old | 81.9 (2.7) | 80.0 (2.8) | 78.5 (3.0) | 76.9 (3.3) | 74.3 (3.8) | -7.6 |
| 1-4 years old | 81.3 (2.6) | 79.5 (2.7) | 78.2 (2.9) | 76.6 (3.2) | 74.1 (3.6) | -7.2 |
| 5-14 years old | 77.4 (2.6) | 75.6 (2.7) | 74.3 (2.9) | 72.7 (3.1) | 70.3 (3.6) | -7.1 |
| 15-24 years old | 67.5 (2.6) | 65.8 (2.7) | 64.4 (2.9) | 62.9 (3.1) | 60.4 (3.6) | -7.1 |
| 25-34 years old | 58.0 (2.6) | 56.2 (2.7) | 54.9 (2.9) | 53.4 (3.1) | 51.0 (3.5) | -7.0 |
| 35-44 years old | 48.5 (2.6) | 46.8 (2.6) | 45.6 (2.8) | 44.1 (3.0) | 41.8 (3.3) | -6.7 |
| 45-54 years old | 38.9 (2.6) | 37.4 (2.6) | 36.3 (2.7) | 35.0 (2.9) | 33.0 (3.1) | -5.9 |
| 55-64 years old | 29.7 (2.5) | 28.5 (2.5) | 27.6 (2.6) | 26.6 (2.6) | 25.0 (2.8) | -4.7 |
| 65-74 years old | 20.9 (2.4) | 20.1 (2.4) | 19.6 (2.4) | 18.9 (2.4) | 17.9 (2.5) | -3.0 |
| 75-84 years old | 13.0 (2.5) | 12.8 (2.4) | 12.6 (2.5) | 12.3 (2.4) | 11.8 (2.5) | -1.2 |

<sup>a</sup> The 85+ age group was excluded since life table estimates in this open interval are generally less precise than in other age groups

The updated ADI (transformed variables) also demonstrated strong associations with age-specific death rates in all age groups (**Figure 2**, **Table 11**). Death rates increased consistently across quintiles of deprivation for all age groups. The relative differences were most pronounced during middle adulthood, with residents of the highest deprivation census tracts experiencing death rates 3.2 times higher than those in the lowest deprivation tracts at ages 45–54.

**Figure 2.**
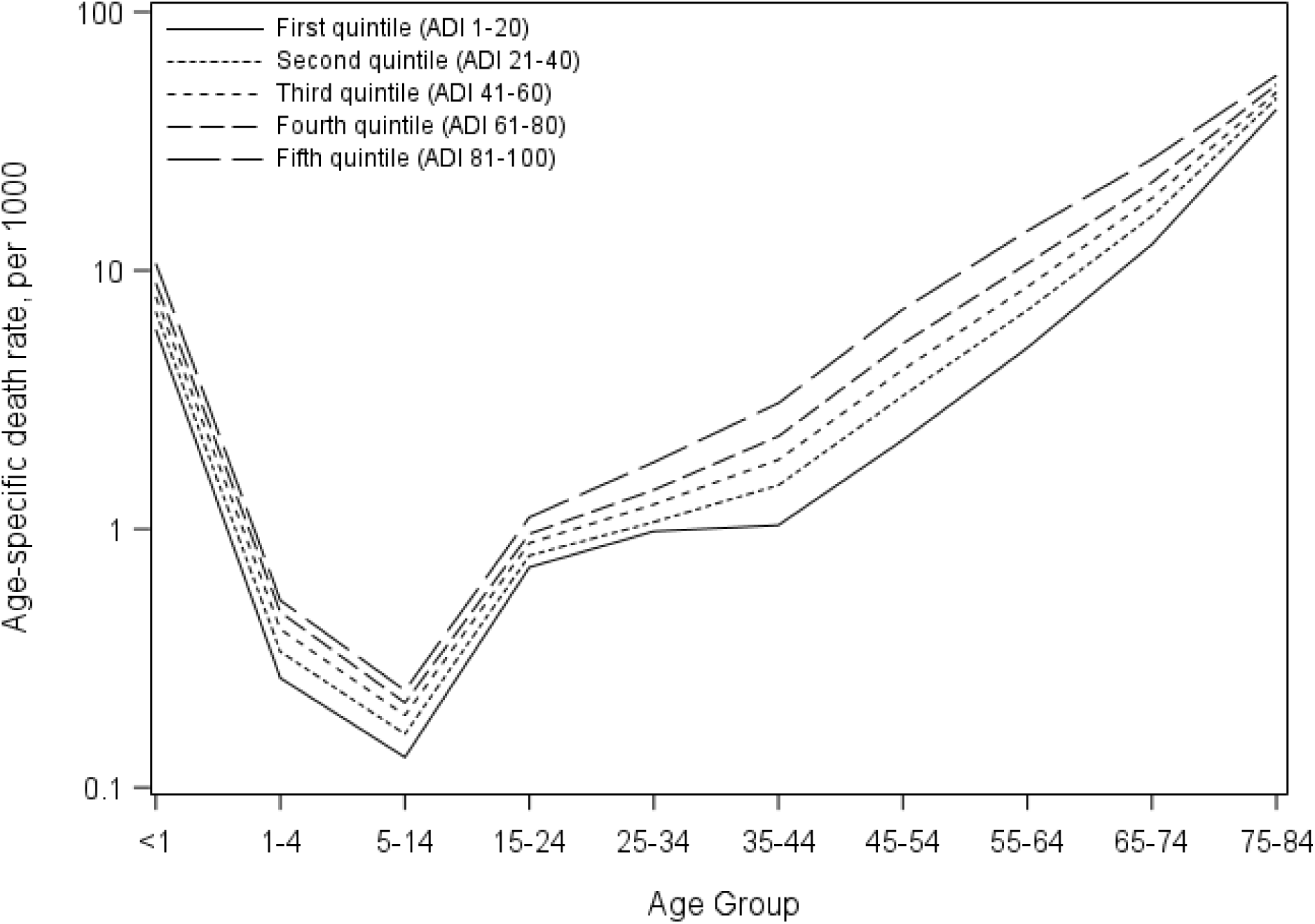
Age-specific death rates by quintile of the updated Area Deprivation Index at the census tract level. The first quintile of ADI (1-20) reflects the census tracts with the least deprivation while the fifth quintile of ADI (81-100) reflects the census tracts with the highest deprivation. *To change: Add “years” to the x-axis labels*

**Table 11.** Mean (SD) tract-level age-specific death rates (ASDR) per 1000 by life table age group and quintiles of ADI.

| Life Table Age Group<sup>a</sup> | Quintile of Duke ADI |  |  |  |  | Ratio of ASDRs<br>Quintile 5 /<br>Quintile 1 |
| --- | --- | --- | --- | --- | --- | --- |
|  | 1 | 2 | 3 | 4 | 5 |  |
| 0-1 years old | 5.9 (4.9) | 6.9 (5.6) | 7.9 (6.3) | 8.9 (7.2) | 10.6 (8.6) | 1.8 |
| 1-4 years old | 0.3 (0.3) | 0.3 (0.4) | 0.4 (0.4) | 0.5 (0.5) | 0.5 (0.5) | 1.7 |
| 5-14 years old | 0.1 (0.2) | 0.2 (0.2) | 0.2 (0.2) | 0.2 (0.2) | 0.2 (0.2) | 2.0 |
| 15-24 years old | 0.7 (0.4) | 0.8 (0.5) | 0.9 (0.5) | 1.0 (0.6) | 1.1 (0.7) | 1.6 |
| 25-34 years old | 1.0 (0.7) | 1.1 (0.7) | 1.2 (0.8) | 1.4 (0.9) | 1.8 (1.2) | 1.8 |
| 35-44 years old | 1.0 (0.6) | 1.5 (0.9) | 1.9 (1.0) | 2.3 (1.3) | 3.1 (1.8) | 3.1 |
| 45-54 years old | 2.2 (1.2) | 3.3 (1.6) | 4.1 (1.8) | 5.2 (2.3) | 7.1 (3.2) | 3.2 |
| 55-64 years old | 5.0 (2.2) | 7.0 (2.8) | 8.6 (3.3) | 10.6 (3.9) | 14.2 (5.6) | 2.8 |
| 65-74 years old | 12.6 (5.0) | 16.2 (5.8) | 19.0 (6.7) | 21.9 (7.6) | 26.9 (10.1) | 2.1 |
| 75-84 years old | 41.9 (15.6) | 46.2 (16.2) | 48.8 (16.6) | 52.2 (17.6) | 56.7 (20.5) | 1.4 |

<sup>a</sup> The 85+ age group was excluded since life table estimates in this open interval are generally less precise than in other age groups

Visual inspection of updated ADI (transformed variables) distributions across census block groups in Washington, DC (**Figure 3**) and Manhattan, New York City (**Figure 4**) provided face validity for the updated index. As previously reported, the Neighborhood Atlas ADI showed considerable uniformity across neighborhoods in both areas.

**Figure 3.**
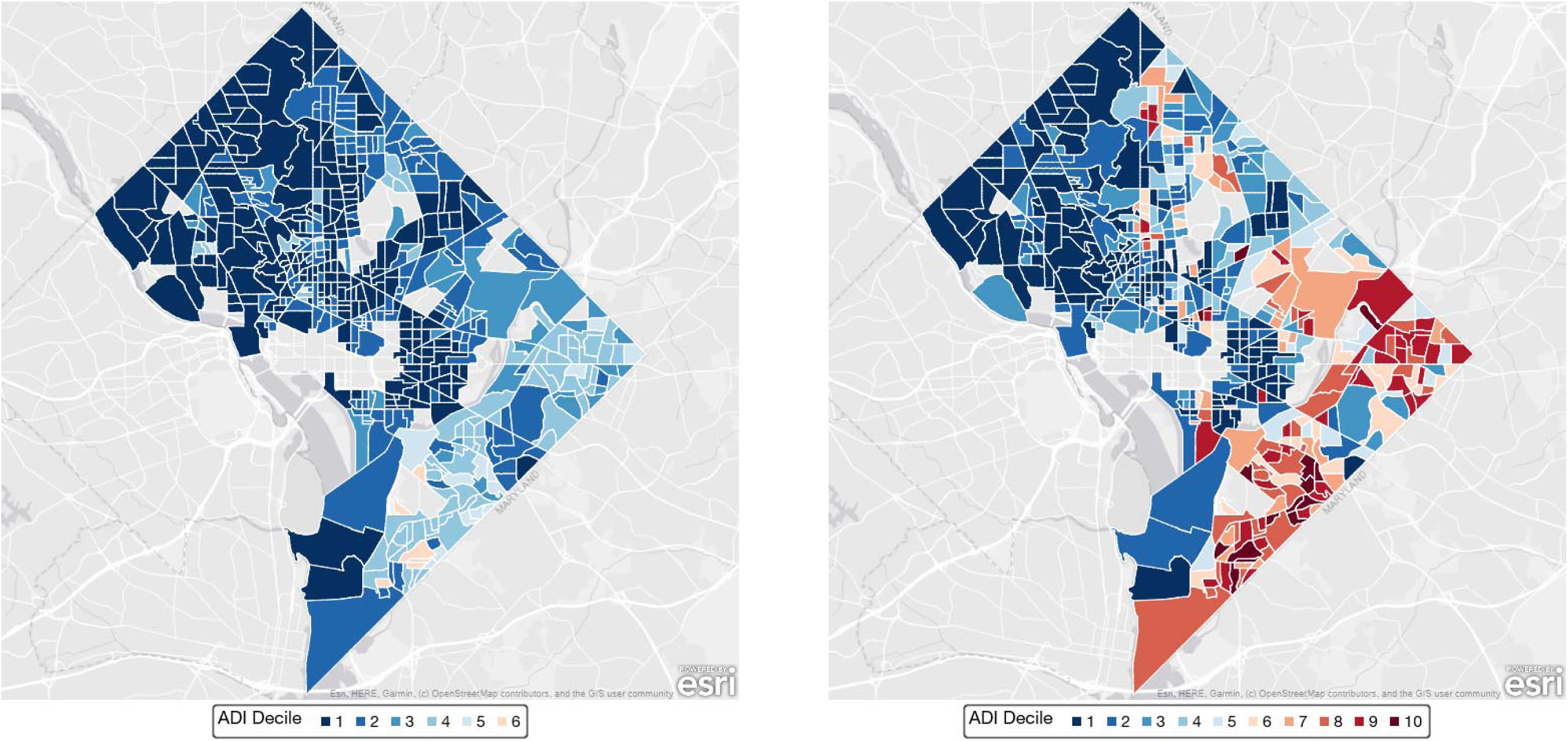
Distribution of the Neighborhood Atlas ADI (left) and the updated ADI (right) by census block groups within Washington, DC. Block groups are shaded by decile of ADI. The first decile (1-10) reflects the block groups with the least deprivation while the tenth decile (91-100) reflects the block groups with the highest deprivation.

**Figure 4.**
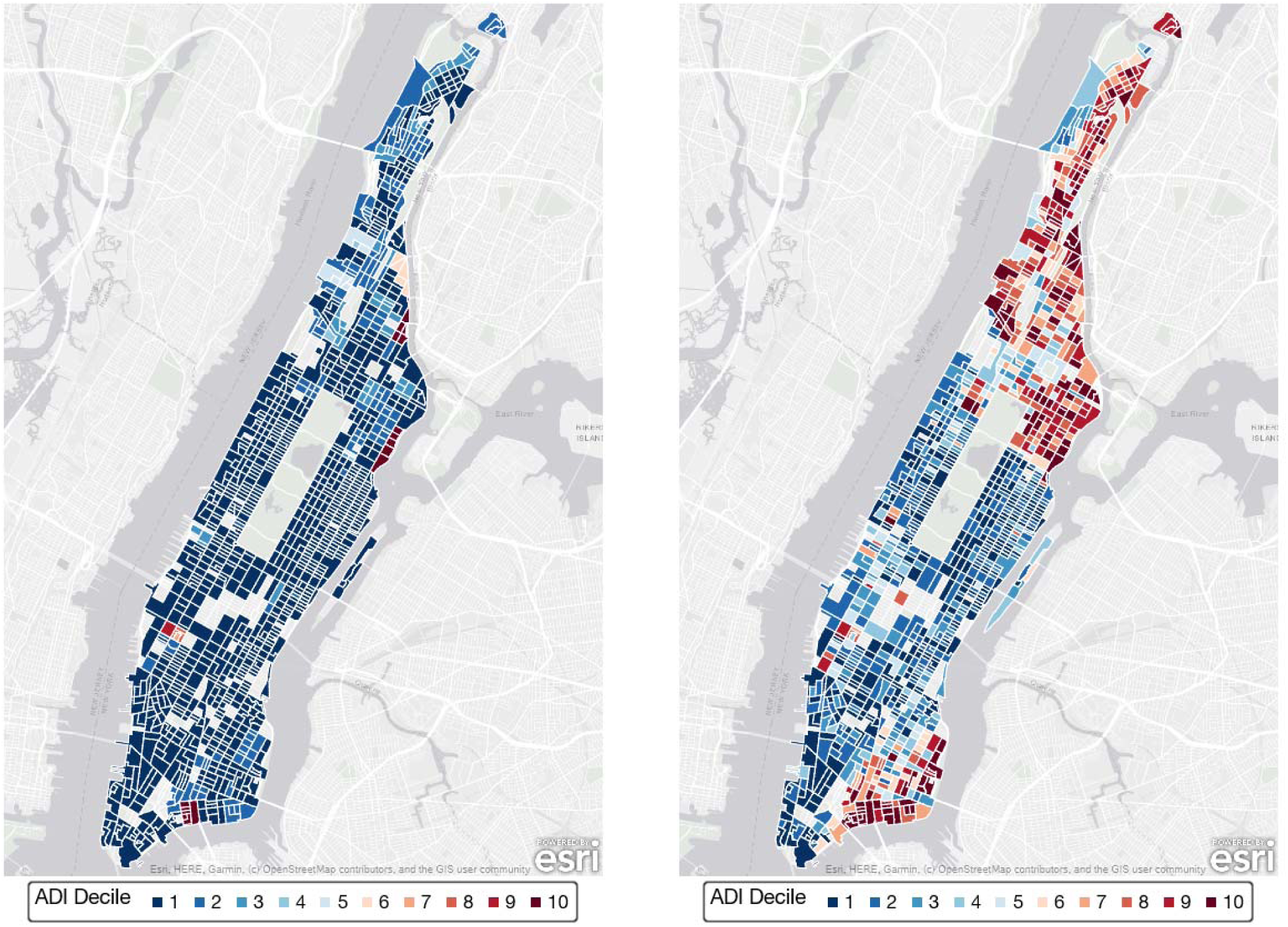
Distribution of the Neighborhood Atlas ADI (left) and the updated ADI (right) by census block groups within Manhattan, New York City. Block groups are shaded by decile of ADI. The first decile (1-10) reflects the block groups with the least deprivation while the tenth decile (91-100) reflects the block groups with the highest deprivation.

## Discussion

In this study, we recalculated the Area Deprivation Index (ADI) using recent American Community Survey (ACS) data after implementing methodologic improvements aimed at strengthening the index’s validity for summarizing contemporary neighborhood socioeconomic conditions. A primary motivation for revisiting the ADI arose from well-documented concerns that the current Neighborhood Atlas version relies heavily on median home value and income, diminishing the contributions of other socioeconomic, housing, employment, and education indicators. In contrast, the updated ADI demonstrated substantial correlations with nearly all 17 component measures, with factor loadings that generally aligned with those from the original ADI developed by Singh using 1990 Census data.

Visual geographic comparisons of ADI values at the block group level in Washington, DC and Manhattan demonstrate the improved face validity of the updated ADI. Both these cities have well-known patterns of socioeconomic heterogeneity that is not reflected by the Neighborhood Atlas ADI. Our updated ADI, on the other hand, identified substantial variation in deprivation that mirror known patterns of inequality.

The updated ADI also showed robust associations with tract-level mortality outcomes. Life expectancy differed by more than seven years between census tracts in the lowest versus highest deprivation quintiles at birth, and meaningful gradients persisted into older adulthood. Age-specific mortality rates followed similar patterns, with the largest relative differences emerging during midlife, where death rates in the highest deprivation quintile were more than three times those of the most advantaged tracts.

Several methodologic recommendations for constructing area-based deprivation measures have been made in recent years, including transparent reporting of analytic processes, updating variable definitions to reflect contemporary conditions, and making source code publicly available. This study directly addresses these critiques. First, by standardizing variables before PCA, we ensure that the resulting index is not disproportionately influenced by variables with inherently larger scales. Second, we modernized the variable set by replacing obsolete measures and updating income disparity thresholds, to reflect substantial increases in U.S. household income distributions since 1990. Third, in line with calls for transparent and reproducible methods, we have made all programming code and updated ADI values publicly available.

The study also highlights areas where future methodological refinement may be warranted. Some measures, such as the percentage of housing units without complete plumbing, were less important in the updated index, suggesting that they may no longer meaningfully capture contemporary deprivation in most communities. As socioeconomic conditions evolve, routine reassessment of the component measures and their operational definitions will be essential. Additionally, while our analyses focused on replicating, updating, and validating the ADI itself, comparative evaluations with social deprivation indices represent an important future research direction.

Taken together, these findings demonstrate that the updated ADI provides a more accurate and comprehensive representation of neighborhood socioeconomic disadvantage than the widely-used Neighborhood Atlas ADI.

## Data Availability

All data produced are available online at https://gitlab.oit.duke.edu/duke-adi/2022/.

## Conflict of Interest

KC: None

BH: None

## Notes

### Competing Interest Statement

The authors have declared no competing interest.

### Funding Statement

This study did not receive any funding.

### Author Declarations

The Duke Health Institutional Review Board approved this study (Pro00106448).

